# LOW PSYCHOLOGICAL WELL-BEING AMONG MEN WHO HAVE SEX WITH MEN (MSM) DURING THE SHELTER-IN-PLACE ORDERS TO PREVENT THE COVID-19 SPREAD: RESULTS FROM A NATIONWIDE STUDY

**DOI:** 10.1101/2020.09.21.20198929

**Authors:** Emerson Lucas Silva Camargo, Bruna Isabela Adolpho de Oliveira, Igor Fessina Siffoni, Anderson Reis de Sousa, Jules Ramon Brito Teixeira, Isabel Amelia Costa Mendes, Alvaro Francisco Lopes de Sousa

## Abstract

**Introduction:** Little is known about how sheltering in place to contain the spread of COVID-19 over extended periods affects individuals’ psychological well-being. This study’s objective was to analyze the factors associated with MSM’s (men who have sex with men) low psychological well-being in the COVID-19 pandemic context.

**Method:** This cross-sectional study was conducted online in the entire Brazilian territory (26 states and federal district) in April and May 2020. The participants were recruited using an adapted version of Respondent-Driven Sampling (RDS), and Facebook posts. Data were collected using social media and MSM dating apps. We estimated the prevalence, crude prevalence ratio (PR), and respective confidence intervals (CI95%).

**Results:** The prevalence of low psychological well-being found in the sample was 7.9%. Associated factors were belonging to the youngest group (PR: 2.76; CI95%: 1.90-4.01), having polyamorous relationships (PR: 2.78; CI95%: 1.51-5.11), not complying with social isolation measures (PR: 6.27; CI95%: 4.42-8.87), not using the social media to find partners (PR: 1.63; CI95%: 1.06-2.53), having multiple sexual partners (PR: 1.80; CI95%: 1.04-3.11), having reduced the number of partners (PR: 2.67; CI95%: 1.44-4.95), and group sex (PR: 1.82; CI95%: 1.23-2.69)

**Conclusion:** The well-being of MSM living in Brazil was negatively affected during the social distancing measures intended to control the spread of COVID-19. The variables that contributed the most to this outcome include social isolation, relationships established with partners, and sexual behavior.

**Policy Implications:** Planning and implementing public policies and actions to promote psychological well-being are needed to improve MSM’s resilience by adopting safe strategies and behavior.

## Introduction

The COVID-19 pandemic is configured as the century’s health challenge. More than 60 million people had been infected worldwide by the end of November 2020 (WHO, 2020), and its impact is apparent in all countries. The populations’ psychosocial health has been considerably affected (Serafini et al., 2020), especially among those more intensively affected by the virus, such as the homeless; migrants and refugees; the elderly; drug users; and indigenous populations (Xiong, et al, 2020). Brazil is currently facing a disheartening situation. It ranks third in the number of COVID-19 cases; by the end of November, it recorded more than 170,000 deaths (Brazil, 2020).

The COVID-19 pandemic context has affected daily life, imposing changes beyond the health sphere, as social, economic, cultural, and political aspects have been impacted with no precedent in the recent history of epidemics (Chakraborty & Maity, 2020). A large number of people infected and dead directly impacts the health systems’ capacity, also challenging the countries’ economy and the populations in general (Araújo, Oliveira & Freitas, 2020). Continuous and prolonged quarantine causes a strain on people’s mental health, as it restricts access to essential goods, such as food, medicines, and transportation, among others (Oliveira et al., 2020). Such a context affects the individuals’ psychological well-being, significantly decreasing quality of life (Serafini et al., 2020). These problems are heightened by mandatory social isolation measures intended to control the virus spread. The damage caused to human well-being is undeniable and has drawn the attention of researchers, lawmakers, health authorities, and public managers, intending to minimize the impact.

The pandemic’s harmful impact, heightened by social isolation, on the individuals’ physical, mental, social, and environmental spheres, is quite noticeable (Forte, Favieri, Tambelli & Casagrande, 2020). In this context, sexual and gender minorities are subject to greater vulnerability, given the historically conditioning inequality they face in terms of health and violence (Gibb et al, 2020). That is, these groups are relegated to a marginal position within the society, and, as a consequence, a safety net system and psychosocial support are not always provided or available to them.

The results reported by a global survey performed by Hornet (Greenhalgh, 2020), a Gay social network, commissioned by the Thomson Reuters Foundation to be conducted among men who have sex with men (MSM), reveal considerable levels of loneliness and depression associated with shelter-in-place measures, emphasizing their broader impact on the individuals’ mental health, which may imply greater impairment in terms of psychological and social well-being throughout the pandemic. The literature has addressed the pandemic’s impact on the maintenance of affective and sexual practices, including the establishment of barriers impeding access to health services, necessary to maintain therapeutic processes and care technologies (Sousa, Oliveira, Schneider, Queiroz, Carvalho, Fronteira, 2020; Carvalho, et al, 2020; Torres et al., 2020). From this perspective, there is a lack of concrete governmental actions directed to MSM during the COVID-19 pandemic.

Given this population’s context, coupled with the repercussions of the novel socio-historical phenomenon caused by the COVID-19 worldwide on the individuals’ psychological well-being, this study’s objective was to analyze the factors associated with low psychological well-being among MSM living in Brazil during the COVID-19 pandemic.

## Methods

### Study design

This cross-sectional study addresses data from the “40TENA” project, conducted in all the 26 Brazilian states and the Federal District from May to April 2020, when restrictive public health measures such as the social isolation and shelter-in-place measures were in force. The Brazilian official health agencies instructed the population to shelter at home, avoiding close/personal contact with people outside one’s household. Essential activities such as trade and some services were kept with restrictions. Up to this study, none of the Brazilian states had imposed full lockdown.

### Population, sample, and eligibility criteria

A total of 2,646 Brazilian MSM participated in this study. Due to the pandemic, the participants were recruited online using Respondent-Driven Sampling (RDS). In this method, the participants themselves are responsible for recruiting other individuals within the same condition via social media. Following the method’s criteria, 15 MSM with different characteristics were selected, namely: the state of residence (we chose the most populated state in each of the five Brazilian regions); Race (Caucasian/non-Caucasian); age (young, adult, or elderly); and educational level (High school, undergraduate or graduate studies). These were the first participants and were called ‘seeds’. Each participant received a link to the survey and was instructed to invite other MSM from their social network until a significant sample was obtained. The seeds were identified through two geo-location-based dating applications (Grindr and Hornet) via direct chat with online users. The first individuals available online in each of the two applications used who met the inclusion criteria were addressed (Queiroz, Sousa et al., 2019; Queiroz et al., 2019 (2); Sousa et al., 2019).

The researchers also boosted the survey on Facebook, directing it to the MSM population aged from 18 to 60 years old (Facebook imposed the age restriction), using a fixed post on the official page of the survey (*https://www.facebook.com/taafimdeque/*) accompanied by an electronic link that granted access to a free and informed consent form and the survey’s questionnaire. Facebook was used as an additional resource due to its ability to access people located in the countryside, extremely necessary in the case of a continental country like Brazil.

Only individuals who identified themselves as men (cisgender or transgender) and aged 18+ years old were included. Tourists and non-Portuguese speaking individuals were excluded.

### Data collection instrument

The instrument used to collect data was developed by this study’s authors, considering the research variables and the participants’ characterization. It was content validated by a panel of judges specialized in the topic and method. The instrument was divided into four sections with 46 questions. Most were multiple-choice questions, and some were mandatory; otherwise, the participant could not proceed with the questionnaire. The questions addressed:

1. Sociodemographic data (age, gender identity, education, sexual orientation, type of relationship, country, state, place of residence);
2. Psychological well-being and coping strategies used during the shelter-in-place orders.

The 5-item World Health Organization Well-Being Index (WHO-5), validated for the Brazilian context (Souza & Hidalgo, 2012), was used to assess psychological well-being. It comprises five items rated on a 5-point Likert scale with a total score ranging from 0 to 25. WHO-5 was designed to measure psychological well-being in the previous two weeks. Scores below 20 indicate the presence of depressive disorder (Souza & Hidalgo, 2012), and scores equal to or lower than 13 indicate impaired well-being (Bech, 2004). Thus, psychological well-being was classified as low (≤13 points), moderate (14 to 19 points), or high (≥20 points).

3 Sexual practice and activities during the pandemic (i.e., casual sex (Carvalho, et al, 2020); sex with the consumption of drugs (Sousa, et al, 2020); use of condoms, protection strategies, strategy used to search for partners, frequency of sexual encounters, and protective measures against COVID-19)
4 Sexual practice and activities before the pandemic (Use of HIV Pre-Exposure Prophylaxis and Post-Exposure Prophylaxis; strategies to search for partners; knowledge regarding STIs and testing), were assessed retrospectively;

Adherence to social distance measures was defined as avoiding personal contact with people outside one’s household for non-essential activities. “Partial” isolation was considered when an individual had direct and recurrent contact with people outside his household, and total isolation when there was no direct contact. The guidelines concerning the presence of signs and symptoms and other clinical variables provided by the Brazilian Ministry of Health were used to define suspect cases of COVID-19 (Brasil, 2020).

For security reasons, the form used to collect data was hosted by a specific site that enabled only one response per IP (internet protocol).

## Data analysis

Data Analysis and Statistical Software (STATA), version 12.0 was used for data analysis. Descriptive and inferential statistics were used. Bivariate analysis was performed using the Chi-square test, estimating prevalence and crude prevalence ratio (PR) with a 95% confidence interval.

Multivariate analysis was performed using Poisson’s Regression analysis with robust variation. The outcome variable (psychological well-being) was included in the analysis, along with each of the associated independent variables with p-value≤0.20. The stepwise procedure was used. The sequence in which each term was inserted in the model was determined by a mutual analysis of theoretical relevance criteria and statistical significance obtained in the bivariate analysis. Each term was added or removed from the model after identifying statistical significance (p-value<0.05), stability of power of association, and Akaike Information Criterion (AIC), defining the useful subset of terms. All the variables with a 5% statistical significance remained in the final model. To determine the best final model, the one with the lowest AIC value was selected.

### Ethical and legal aspects

The research project was approved by the Institutional Review Board at the Universidade Nova de Lisboa and the University of São Paulo. All the users signed free and informed consent forms before proceeding with the questionnaire.

## Results

A total of 2,646 men who have sex with men participated, 1,921 (72.6%) of whom were referred by their partners or friends from other social networks. Most men presented moderate psychological well-being (47.7%), followed by those with high well-being (44.4%), while a prevalence ratio of 7.9% experiencing low well-being was found. Most were aged from 18 to 29 years old (69.9%), 31 years old on average (Min. 18 and Max. 60; SD±8.4), single (67.2%), fully complying with shelter-in-place orders (72.1%), with a duration of social isolation between 30 and 45 days (58.1%), and decreased consumption of alcohol (43.1%) (Table 1).

**Table 1.**
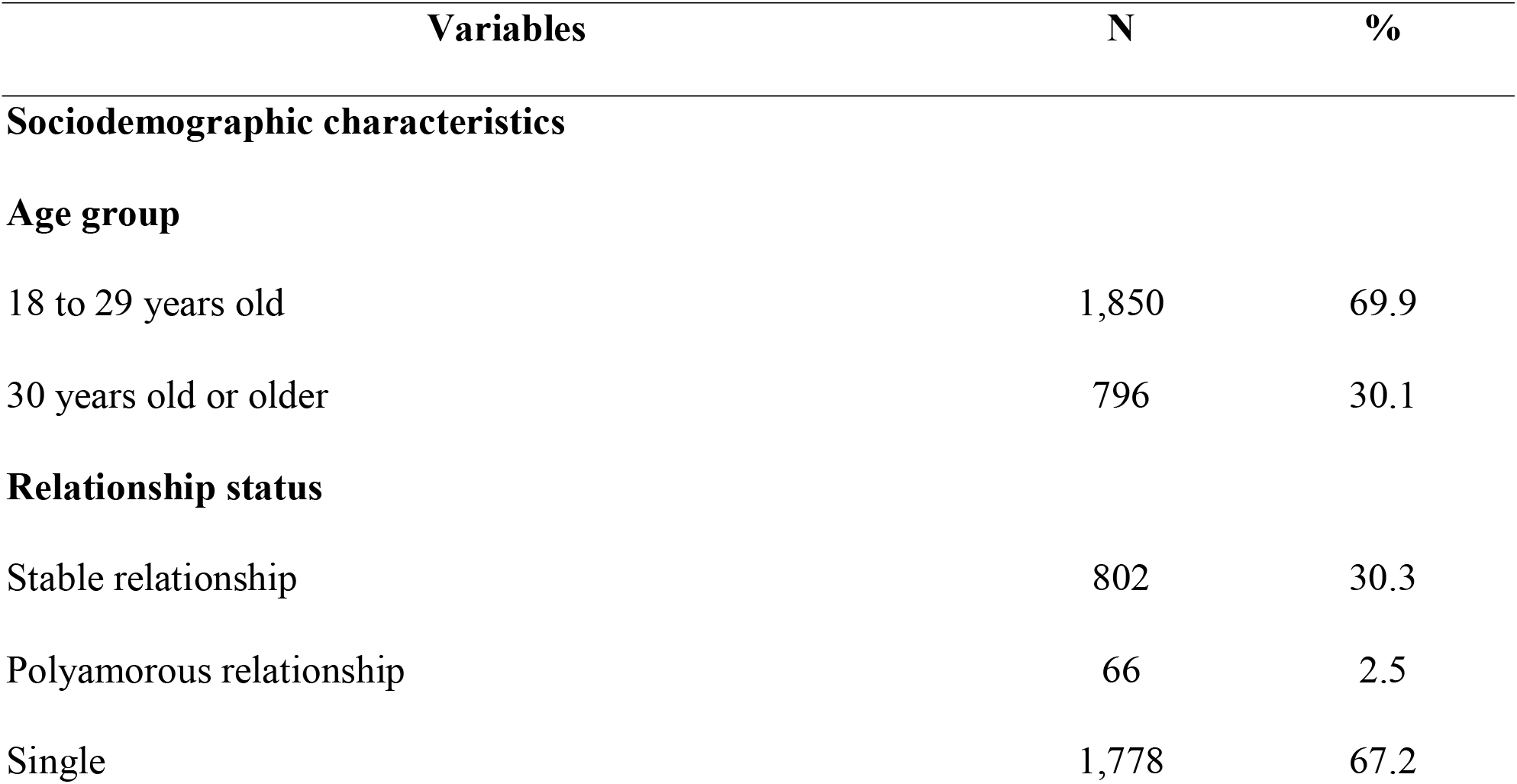
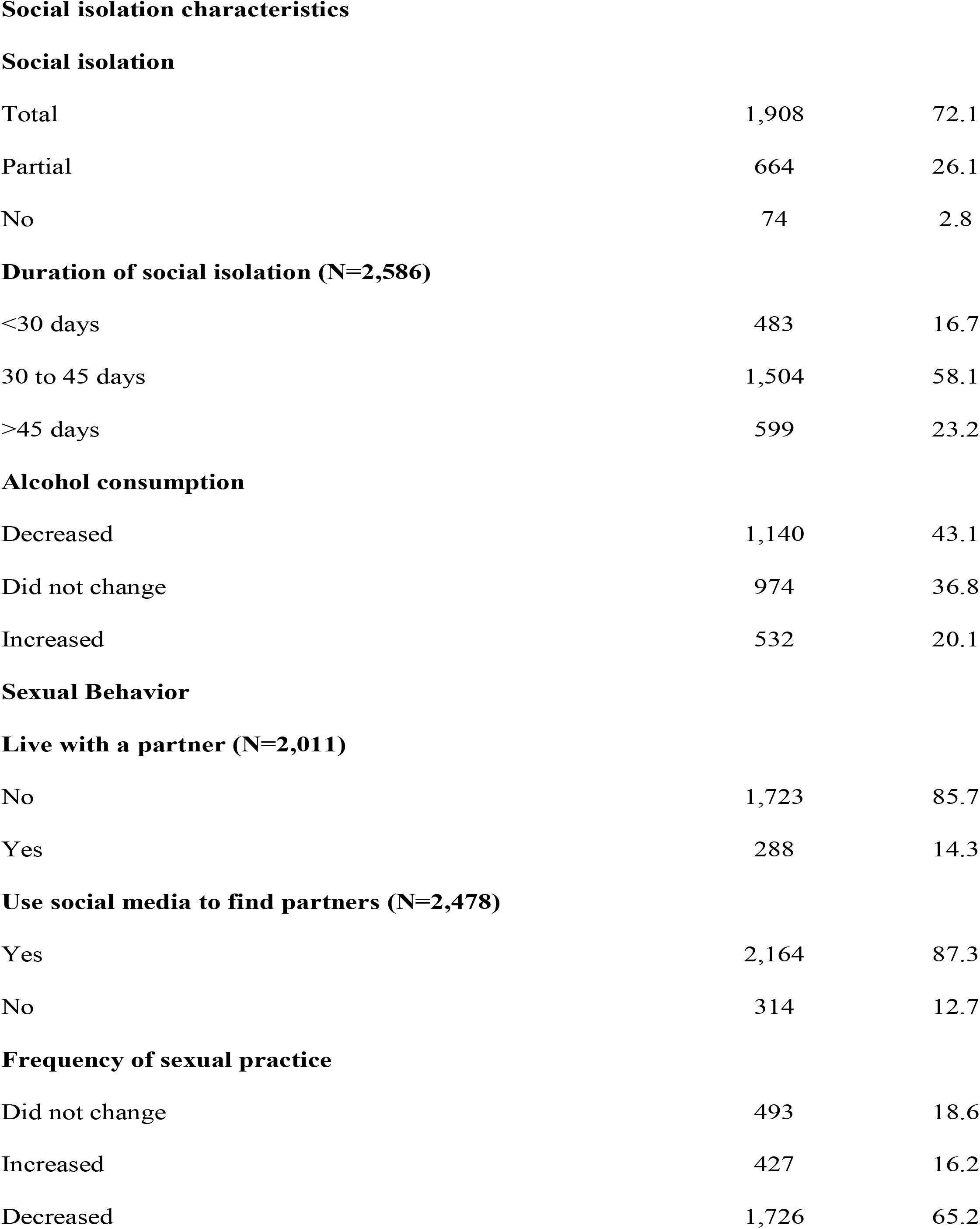
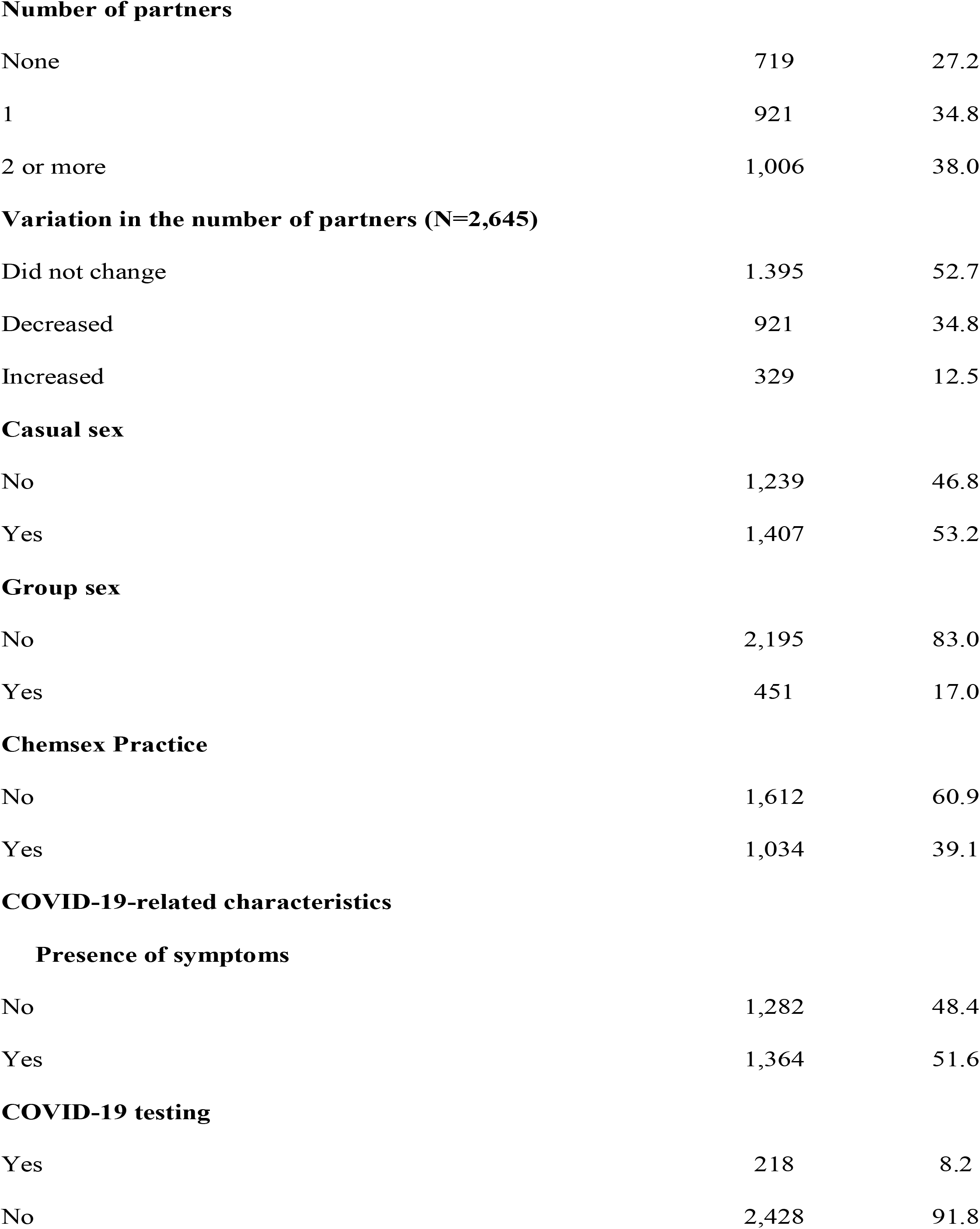
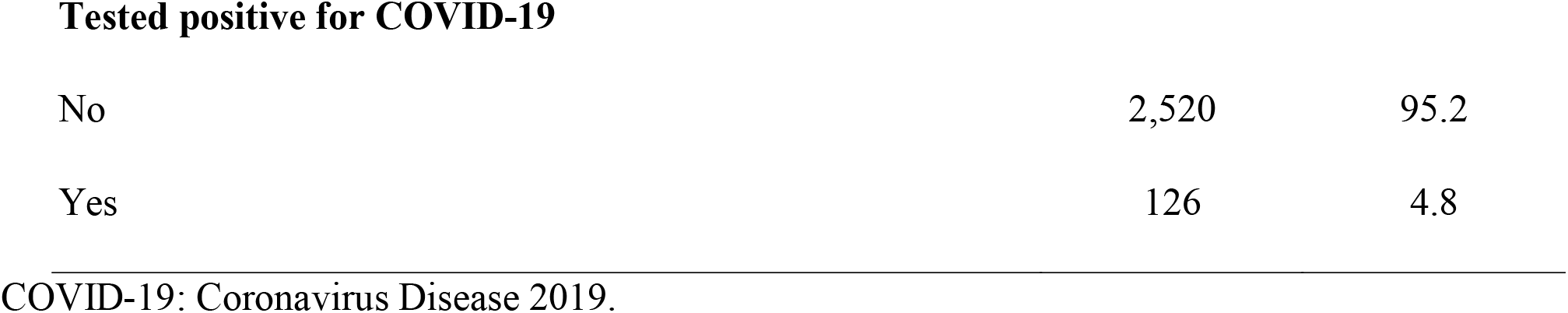
Distribution of men according to sociodemographic characteristics, adherence to social isolation measures, sexual behavior, and COVID-19-related characteristics. Brazil, 2020. (N=2,646).

| Variables | N | % |
| --- | --- | --- |
| **Sociodemographic characteristics** |  |  |
| **Age group** |  |  |
| 18 to 29 years old | 1,850 | 69.9 |
| 30 years old or older | 796 | 30.1 |
| **Relationship status** |  |  |
| Stable relationship | 802 | 30.3 |
| Polyamorous relationship | 66 | 2.5 |
| Single | 1,778 | 67.2 |
| **Social isolation characteristics****Social isolation** |  |  |
| Total | 1,908 | 72.1 |
| Partial | 664 | 26.1 |
| No | 74 | 2.8 |
| **Duration of social isolation (N=2,586)** |  |  |
| <30 days | 483 | 16.7 |
| 30 to 45 days | 1,504 | 58.1 |
| >45 days | 599 | 23.2 |
| **Alcohol consumption** |  |  |
| Decreased | 1,140 | 43.1 |
| Did not change | 974 | 36.8 |
| Increased | 532 | 20.1 |
| **Sexual Behavior****Live with a partner (N=2,011)** |  |  |
| No | 1,723 | 85.7 |
| Yes | 288 | 14.3 |
| **Use social media to find partners (N=2,478)** |  |  |
| Yes | 2,164 | 87.3 |
| No | 314 | 12.7 |
| **Frequency of sexual practice** |  |  |
| Did not change | 493 | 18.6 |
| Increased | 427 | 16.2 |
| Decreased | 1,726 | 65.2 |
| **Number of partners** |  |  |
| None | 719 | 27.2 |
| 1 | 921 | 34.8 |
| 2 or more | 1,006 | 38.0 |
| **Variation in the number of partners (N=2,645)** |  |  |
| Did not change | 1,395 | 52.7 |
| Decreased | 921 | 34.8 |
| Increased | 329 | 12.5 |
| **Casual sex** |  |  |
| No | 1,239 | 46.8 |
| Yes | 1,407 | 53.2 |
| **Group sex** |  |  |
| No | 2,195 | 83.0 |
| Yes | 451 | 17.0 |
| **Chemsex Practice** |  |  |
| No | 1,612 | 60.9 |
| Yes | 1,034 | 39.1 |
| **COVID-19-related characteristics****Presence of symptoms** |  |  |
| No | 1,282 | 48.4 |
| Yes | 1,364 | 51.6 |
| **COVID-19 testing** |  |  |
| Yes | 218 | 8.2 |
| No | 2,428 | 91.8 |
| Tested positive for COVID-19 |  |  |
| No | 2,520 | 95.2 |
| Yes | 126 | 4.8 |

COVID-19: Coronavirus Disease 2019.

The bivariate analysis of factors associated with psychological well-being showed statistical association with age (p<0.001), relationship status (p<0.001), and social isolation (p<0.001).

Being 18 to 29 years old, in a polyamorous relationship, and not complying with shelter-in-place orders increased three, twice, and seven times, respectively, the prevalence of low psychological well-being. Social isolation increased the frequency of low psychological well-being by 47%. Other findings are presented in Table 2.

**Table 2.**
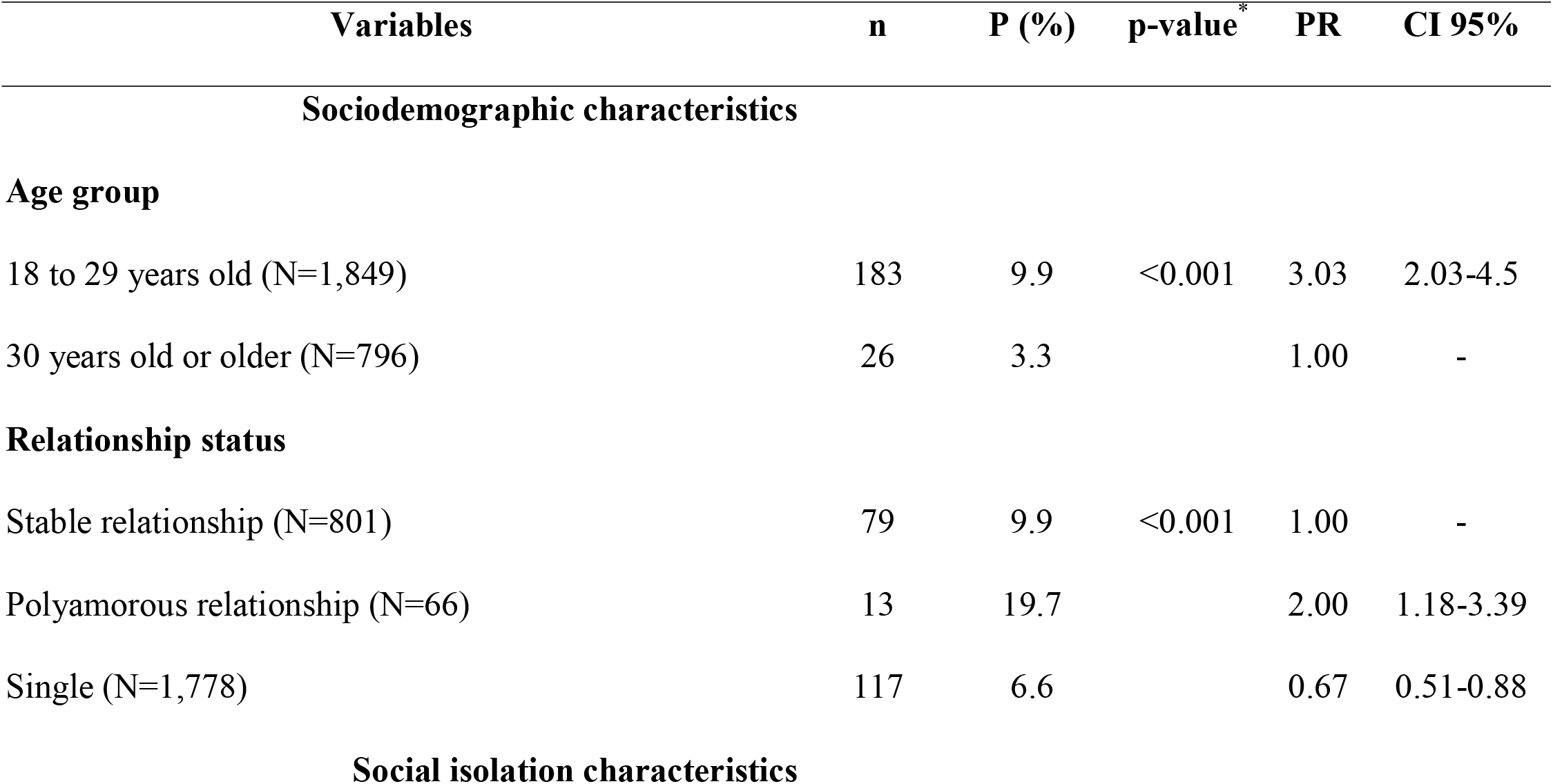
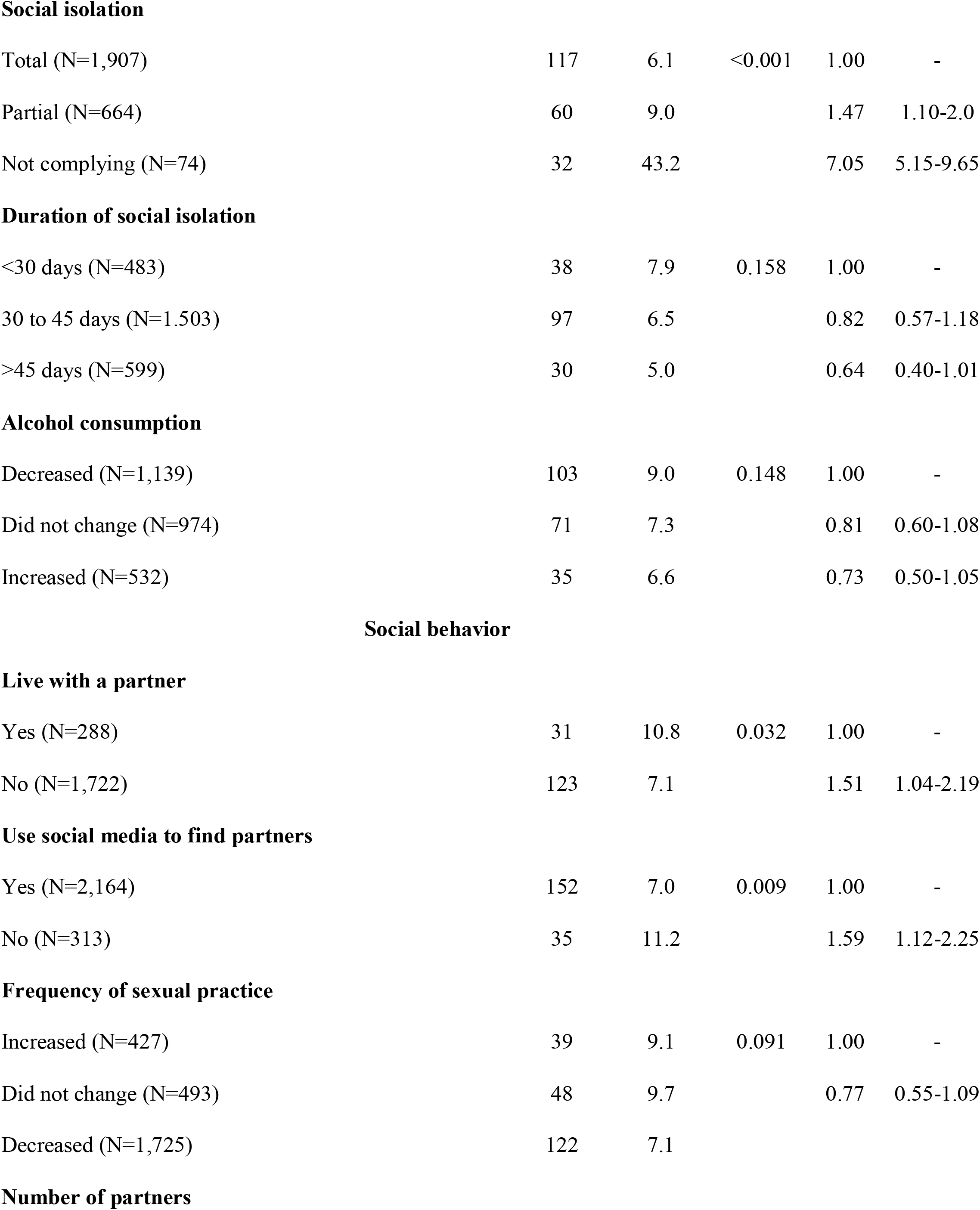
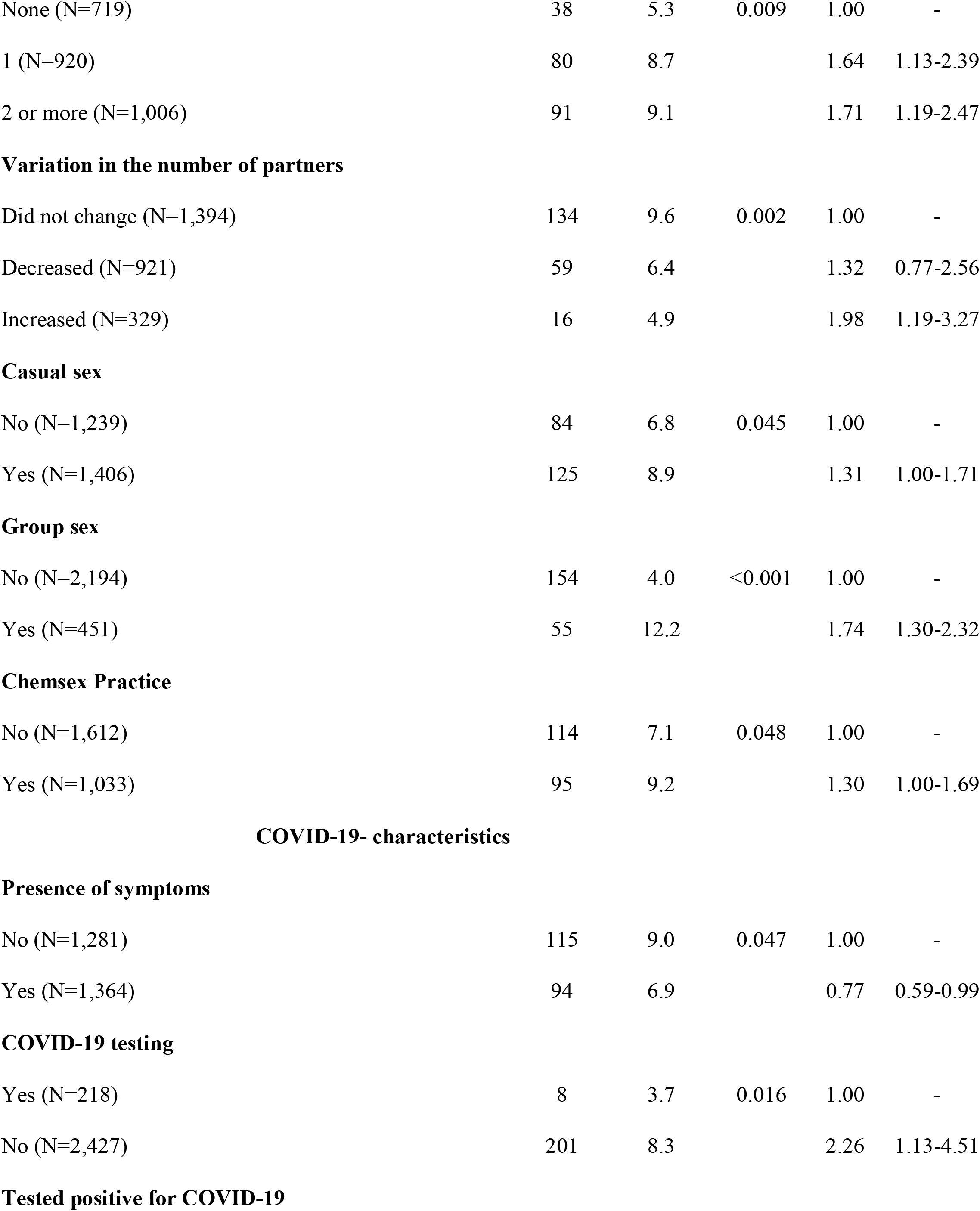
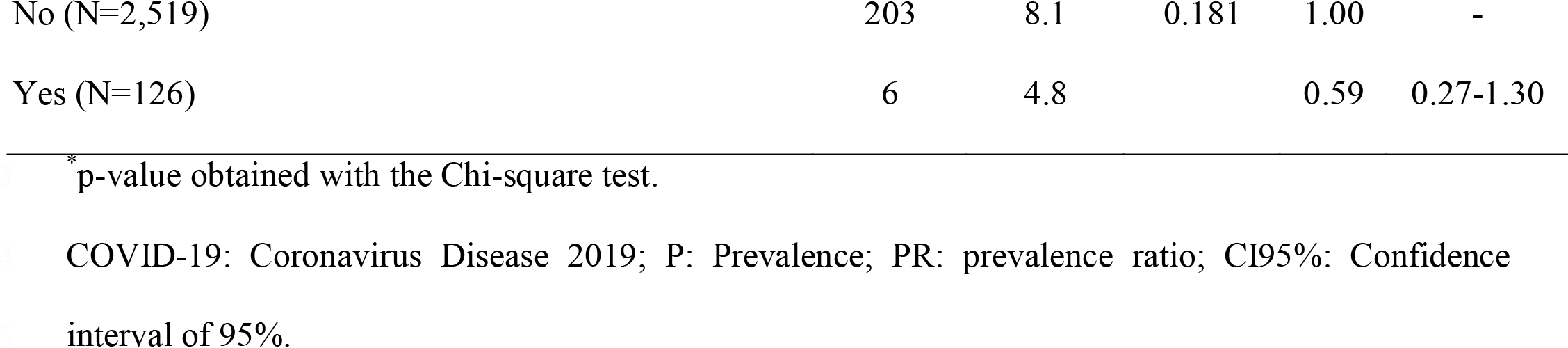
Bivariate analysis between psychological well-being and sociodemographic characteristics, adherence to social isolation, sexual behavior, and COVID-19-related characteristics. Brazil, 2020.

| Variables | n | P (%) | p-value<sup>*</sup> | PR | CI 95% |
| --- | --- | --- | --- | --- | --- |
| **Sociodemographic characteristics** |  |  |  |  |  |
| **Age group** |  |  |  |  |  |
| 18 to 29 years old (N=1,849) | 183 | 9.9 | <0.001 | 3.03 | 2.03-4.5 |
| 30 years old or older (N=796) | 26 | 3.3 |  | 1.00 | - |
| **Relationship status** |  |  |  |  |  |
| Stable relationship (N=801) | 79 | 9.9 | <0.001 | 1.00 | - |
| Polyamorous relationship (N=66) | 13 | 19.7 |  | 2.00 | 1.18-3.39 |
| Single (N=1,778) | 117 | 6.6 |  | 0.67 | 0.51-0.88 |
| **Social isolation characteristics** |  |  |  |  |  |
| **Social isolation** |  |  |  |  |  |
| Total (N=1,907) | 117 | 6.1 | <0.001 | 1.00 | - |
| Partial (N=664) | 60 | 9.0 |  | 1.47 | 1.10-2.0 |
| Not complying (N=74) | 32 | 43.2 |  | 7.05 | 5.15-9.65 |
| **Duration of social isolation** |  |  |  |  |  |
| <30 days (N=483) | 38 | 7.9 | 0.158 | 1.00 | - |
| 30 to 45 days (N=1,503) | 97 | 6.5 |  | 0.82 | 0.57-1.18 |
| >45 days (N=599) | 30 | 5.0 |  | 0.64 | 0.40-1.01 |
| **Alcohol consumption** |  |  |  |  |  |
| Decreased (N=1,139) | 103 | 9.0 | 0.148 | 1.00 | - |
| Did not change (N=974) | 71 | 7.3 |  | 0.81 | 0.60-1.08 |
| Increased (N=532) | 35 | 6.6 |  | 0.73 | 0.50-1.05 |
| **Social behavior****Live with a partner** |  |  |  |  |  |
| Yes (N=288) | 31 | 10.8 | 0.032 | 1.00 | - |
| No (N=1,722) | 123 | 7.1 |  | 1.51 | 1.04-2.19 |
| **Use social media to find partners** |  |  |  |  |  |
| Yes (N=2,164) | 152 | 7.0 | 0.009 | 1.00 | - |
| No (N=313) | 35 | 11.2 |  | 1.59 | 1.12-2.25 |
| **Frequency of sexual practice** |  |  |  |  |  |
| Increased (N=427) | 39 | 9.1 | 0.091 | 1.00 | - |
| Did not change (N=493) | 48 | 9.7 |  | 0.77 | 0.55-1.09 |
| Decreased (N=1,725) | 122 | 7.1 |  |  |  |
| **Number of partners** |  |  |  |  |  |
| None (N=719) | 38 | 5.3 | 0.009 | 1.00 | - |
| 1 (N=920) | 80 | 8.7 |  | 1.64 | 1.13-2.39 |
| 2 or more (N=1,006) | 91 | 9.1 |  | 1.71 | 1.19-2.47 |
| **Variation in the number of partners** |  |  |  |  |  |
| Did not change (N=1,394) | 134 | 9.6 | 0.002 | 1.00 | - |
| Decreased (N=921) | 59 | 6.4 |  | 1.32 | 0.77-2.56 |
| Increased (N=329) | 16 | 4.9 |  | 1.98 | 1.19-3.27 |
| **Casual sex** |  |  |  |  |  |
| No (N=1,239) | 84 | 6.8 | 0.045 | 1.00 | - |
| Yes (N=1,406) | 125 | 8.9 |  | 1.31 | 1.00-1.71 |
| **Group sex** |  |  |  |  |  |
| No (N=2,194) | 154 | 4.0 | <0.001 | 1.00 | - |
| Yes (N=451) | 55 | 12.2 |  | 1.74 | 1.30-2.32 |
| **Chemsex Practice** |  |  |  |  |  |
| No (N=1,612) | 114 | 7.1 | 0.048 | 1.00 | - |
| Yes (N=1,033) | 95 | 9.2 |  | 1.30 | 1.00-1.69 |
| **Presence of symptoms** |  |  |  |  |  |
| No (N=1,281) | 115 | 9.0 | 0.047 | 1.00 | - |
| Yes (N=1,364) | 94 | 6.9 |  | 0.77 | 0.59-0.99 |
| **COVID-19 testing** |  |  |  |  |  |
| Yes (N=218) | 8 | 3.7 | 0.016 | 1.00 | - |
| No (N=2,427) | 201 | 8.3 |  | 2.26 | 1.13-4.51 |
| **Tested positive for COVID-19** |  |  |  |  |  |
| No (N=2,519) | 203 | 8.1 | 0.181 | 1.00 | - |
| Yes (N=126) | 6 | 4.8 |  | 0.59 | 0.27-1.30 |

\*p-value obtained with the Chi-square test.
COVID-19: Coronavirus Disease 2019; P: Prevalence; PR: prevalence ratio; CI95%: Confidence interval of 95%.

The variable ‘type of partner’ was excluded from the multivariate analysis because it presented collinearity with relationship status (VIF=7.37). The association between the presence of COVID-19 symptoms and testing for COVID-19 was not statistically significant but remained in the model as confounding adjustment because the model presented a lower AIC.

The following variables were associated with low psychological well-being: being in the youngest age group (PR: 2.76; CI95%: 1.90-4.01); having a polyamorous relationship (PR: 2.78; CI95%: 1.51-5.11); not complying with social isolation measures (PR: 6.27; CI95%: 4.42-8.87); not using the social media to find partners (PR: 1.63; CI95%: 1.06-2.53); having a higher number of sexual partners (PR: 1.80; CI95%: 1.04-3.11); having reduced the number of sexual partners (PR: 2.67; CI95%: 1.44-4.95); and group sex (PR: 1.82; CI95%: 1.23-2.69) (Table 3).

**Table 3.**
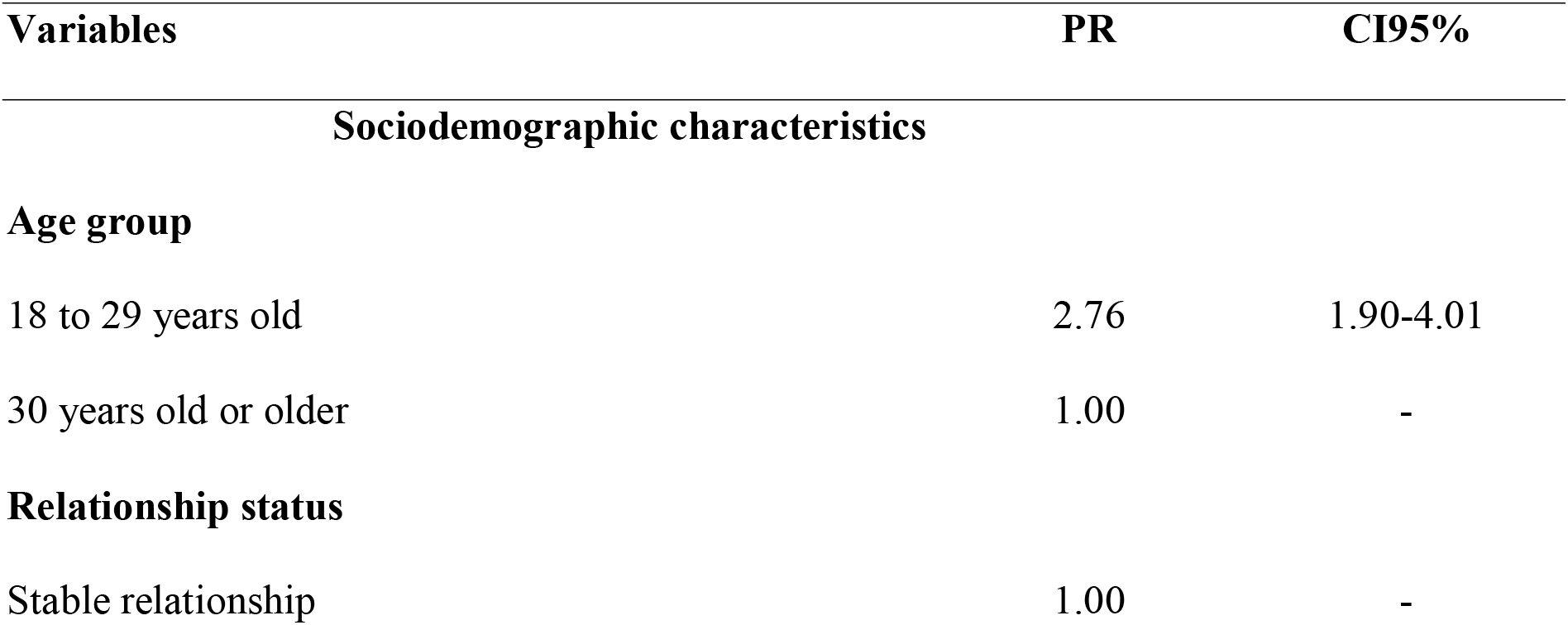
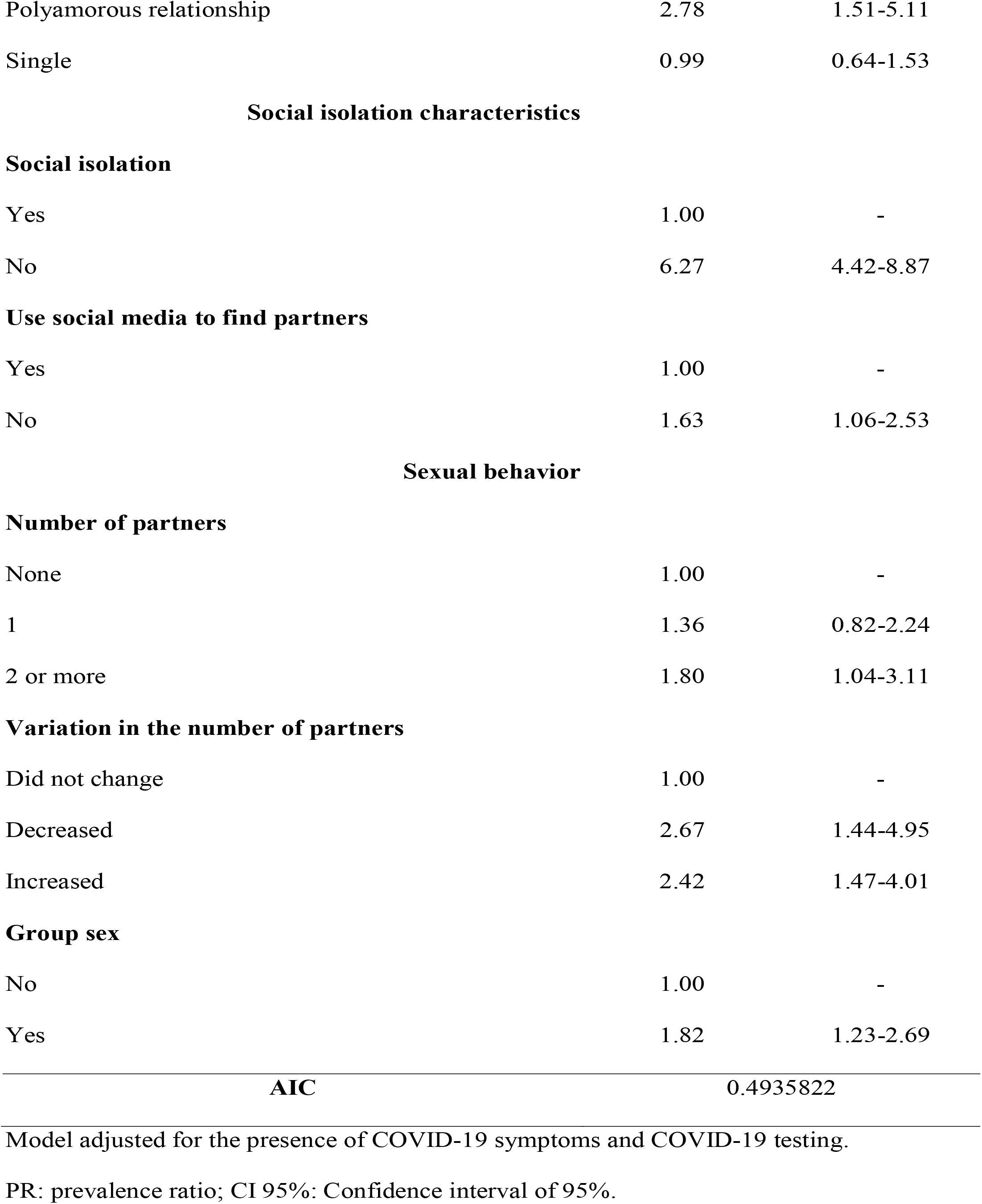
Multivariate analysis of factors associated with psychological well-being. Brazil, 2020.

| Variables | PR | CI95% |
| --- | --- | --- |
| **Sociodemographic characteristics** |  |  |
| **Age group** |  |  |
| 18 to 29 years old | 2.76 | 1.90-4.01 |
| 30 years old or older | 1.00 | - |
| **Relationship status** |  |  |
| Stable relationship | 1.00 | - |
| Polyamorous relationship | 2.78 | 1.51-5.11 |
| Single | 0.99 | 0.64-1.53 |
| **Social isolation** |  |  |
| Yes | 1.00 | - |
| No | 6.27 | 4.42-8.87 |
| **Use social media to find partners** |  |  |
| Yes | 1.00 | - |
| No | 1.63 | 1.06-2.53 |
| **Number of partners** |  |  |
| None | 1.00 | - |
| 1 | 1.36 | 0.82-2.24 |
| 2 or more | 1.80 | 1.04-3.11 |
| **Variation in the number of partners** |  |  |
| Did not change | 1.00 | - |
| Decreased | 2.67 | 1.44-4.95 |
| Increased | 2.42 | 1.47-4.01 |
| **Group sex** |  |  |
| No | 1.00 | - |
| Yes | 1.82 | 1.23-2.69 |

## Discussion

MSM living in Brazil experienced worse well-being during the shelter-in-place orders implemented to restrain the spread of COVID-19, as more than half of the participants presented some level of impairment in terms of psychological well-being. The variables that most frequently contributed to this outcome include social isolation/distancing and variables related to the type of relationship and sexual behavior (not complying with social isolation measures, strategies used to search for sexual partners, number of sexual partners, and group sex, stood out).

The declining indicators of psychological well-being among MSM are worth noting. In a devastating context, such as that of a pandemic, access to psychosocial support networks and the individuals’ socio-affective network is considerably weakened and even absent in some instances, potentially intensifying these individuals’ vulnerability from the perspective of mental health and social well-being. For instance, the sudden rupture in socio-affective networks caused by the COVID-19 pandemic, not having access to nightlife or leisure, not meeting friends and groups of belonging, not having intimate and sexual encounters, not being able to keep affective relationships, and interactions being restricted to the virtual environment or small groups, may be quite challenging.

For this reason, the current pandemic context requires we consider the quality or lack of social interactions and how these situations impact the psychological well-being of people as psychological triggers, such as loneliness, change in routine, abandonment, fear, loss of financial stability, or unemployment, may be intensified (SCHMIDT et al., 2020).

This context is likely to cause anxiety, decision conflicts, decreased self-esteem and interest in life, suicidal behavior, eating and sleep disorders, and abusive alcohol or drug consumption among MSM (Pedrosa et al., 2020; Leung et al., 2020). These individuals may even become involved in practices in which they expose themselves to the Coronavirus for not bearing the prolonged social distancing measures, boredom, and the lack of social interaction with people, or restricted mobility within the cities (Reis de Sousa et al., 2020). Coupled with these is the fact that difficulty in searching for affective and sexual partners during the period of social isolation may also decrease psychological well-being, impacting the health of these men (Sousa & Oliveira et al, 2020; Carvalho et al., 2020).

In this study, a greater prevalence of low psychological well-being was found among MSM who did not report the use of social media to find partners, revealing that even though the social media is considered to be associated with sex, in times of social distancing, it may play a protective role by enabling social interactions, even if remotely. The results from a multi-center study show that 95% of its MSM sample was (totally or partially) complying with social distancing measures, which resulted in a drop in the number of partners and relationships, but, at the same time, increased online interactions (Sousa & Oliveira et al., 2020). The study also reports that MSM devised strategies to keep interactions during the social distancing measures, reinforcing this study’s findings that interactions, whether to seek or exchange content, are essential for MSM’s psychological well-being. The study mentioned above addressed only sexual interactions though, so inferences concerning other types of relationships are impossible.

Interestingly, having a polyamorous relationship (three people or more) was associated with low psychological well-being. One hypothesis is that polyamorous relationships do not always occur under the same roof, implying that those involved may have to deal with distancing measures; that is, they may not contact each other for some time. Additionally, polyamorous relationships allow new people, outside one’s place of shelter and with a possibly different history of exposure, to be included. In the case of physical contact, all those involved are exposed, which results in fear, and decreased psychological well-being.

Consensual non-monogamy or polyamorous relationships have become an increasingly accepted affective and sexual experience, which does not comply with monogamous relationship patterns, usually considered ideal in stable and committed relationships (Costa & Ribeiro, 2020; Jowett, 2020). This context has been intensified among sexual minorities, including MSM. We believe that the fact these individuals are involved in polyamorous relationships and are currently experiencing the impossibility to maintain the frequency of affective and sexual meetings during the pandemic, that is, impaired affective and sexual dynamics are coupled with diminished leisure opportunities these relationships usually promote, decreasing these men’s psychological well-being.

One cannot rule out the fact that MSM in polyamorous relationships have to live with stigma. In the context of this pandemic, this experience may be heightened considering the disruptive phenomena caused by the stigmatization process such as labels, judgment, withdrawing, and discredit, negatively impacting their physical and mental health, and consequently their psychological well-being (Conley, Piemonte, Gusakova & Rubin, 2018; Conley, 2017).

In line with this hypothesis, this study shows that having a higher number of sexual partners and practicing group sex was associated with low psychological well-being, which reinforces the hypothesis that having sex with casual partners implies exposure that includes the risk of acquiring sexually transmissible infections and COVID-19 (Stephenson et al, 2020). Sex is a pleasurable practice to satisfy one’s desire and improves an individual’s general well-being. Group sex, which is usually a source of satisfaction, in a pandemic context, can lead to psychological distress afterward due to a fear of having being infected by COVID-19 (Sousa et al., 2020). Although MSM’s sexual practices incur risks, we have to bear in mind that such practices are diverse and vary in terms of configurations and relationships, requiring a positive approach considering the specificities of each MSM group.

Exposure to often unknown partners, or familiar partners who do not effectively adopt preventive measures, including those against COVID-19, increases the fear of being contaminated or anxiety with the possibility of presenting signs and symptoms of the disease, leading to low psychological well-being (Newman & Guta, 2020; Suen, Chan & Wong, 2020; Brennan, Card, Collict, Jolimore & Lachowsky, 2020).

Considering the disciplinary role of affective and sexual practices, which coupled with the advent of a novel infectious disease, for which an effective treatment or cure has not been devised thus far, keeping multiple sexual partners and/or having group sex may increase the levels of anxiety, stress, uncertainty, worry, and fear among MSM. Additionally, the COVID-19 epidemiological curve in Brazil at the time of data collection was ascending, with an expressive number of new cases and deaths (Silva et al., 2020). Concomitantly, there was an increase in the amount of information provided by the media, exposing the complexity of the disease and virulence of SARS-CoV-2, which also tends to influence the individuals’ psychological well-being negatively (Gao et al, 2020).

The prevalence of low psychological well-being was approximately six times greater among MSM who did not comply or could not comply with the social isolation measures, reinforcing the hypothesis that even though the individuals may not comply with social distancing measures to seek interactions, exposure still may cause concerns (Sousa & Oliveira et al., 2020; Newman & Guta, 2020; This finding should be interpreted with caution because this study did not assess the context in which people violated social isolation measures, socialized with roommates, family, and co-workers, or yet how people interpreted the questions. Additionally, we should consider that seeking social interactions might not be a matter of choice but rather a compulsive behavior, even though it implies decreased psychological well-being afterward (Sanchez et al, 2020).

Nonetheless, having a small number of partners also affected these individuals’ psychological well-being negatively. If, on the one hand, the individuals want to decrease exposure to protect themselves, on the other hand, not having the social support provided by relationships and partners, which would decrease the pandemic’s harmful effects, could negatively affect these individuals’ mental health (Sanchez et al., 2020; Carrico, et al., 2020; Newman & Guta, 2020).

Variation in the number of affective and sexual partners may have influenced and decreased the psychological well-being of MSM and is associated with the low psychosocial support enabled by the measures imposed by Brazilian agencies to protect people or minimize the impact of the pandemic context on mental health. Longitudinal studies are needed to investigate this finding in more detail. In Brazil, stress has been intensified among minority social groups as their ‘invisibility’ has been exposed along with denial of rights, cuts on budgets previously designated to the LGBTQIA+ population, and naturalized institutional homophobia (Folha de São Paulo, 2020).

A lack of directive public policies mainly affects the youngest population, which in this study presented the highest prevalence of low psychological well-being, almost three times higher than among older MSM. The World Health Organization has shown concern with the youngest population within the context of the COVID-19 pandemic, mainly because this is the most vulnerable in not adhering to social isolation measures. An increase in the number of new COVID-19 cases, agglomerations, and non-adherence to public health measures to control the spread of Sars-CoV-2, has been found in countries already in the summer season (Young mind, 2020; Cohen & Bosk, 2020).

The Inter-Agency Standing Committee (IASC) has proposed specific guidelines to promote mental health and strengthen psychosocial support in emergencies, as is the case of the COVID-19 pandemic. These guidelines focus on promoting people’s psychological well-being and preventing and/or treating mental diseases (IASC, 2007). These goals are achieved by employing multiple approaches that take into account biological, sociocultural, educational, and community characteristics, which should focus on reducing harm, promoting human rights and equality, social participation, and valorization of local resources and competencies, the work via integrated support systems, and rehabilitation (Dong, Du, & Gardner, 2020; Fukuti et al., 2020).

Socio-affective networks are a significant source of access to protection, sociability, exchange, and maintenance of MSM’s psychological well-being and health. In the pandemic context, and even after the pandemic, supporting, strengthening, and disseminating these networks may be an effective strategy to improve the levels of psychological well-being, decreasing the impact caused by social isolation, such as feelings of loneliness, heightened anxiety, and stress.

This study’s limitations include its design. Cross-sectional studies do not allow for cause and effect conclusions; that is, longitudinal studies would allow for better evidence of associations. Additionally, due to the need to comply with social distancing measures, population-based face-to-face interviews were not possible, except for the MSM who did not have access to the Internet and could not complete the electronic form. Finally, a probabilistic sampling procedure was not adopted here, which impedes the generalization of results to other contexts or countries, though the results provide a situational diagnosis of the MSM’s psychological well-being in the pandemic context.

## Conclusion

The factors associated with the MSM’s low psychological well-being were: belonging to the youngest age group; being in a polyamorous relationship; not complying with the social isolation measures; not using social media to find partners; having multiple sexual partners; having reduced the number of partners, and group sex.

The planning and implementation of public policies and actions to promote psychological well-being and mental health in general, intended to increase resilience among MSM are needed. Safe sexual strategies and behavior need to be promoted, along with the use of digital technologies to facilitate social relationships by enabling remote interactions. These measures can alleviate the harmful effects of the intensified psychological needs accruing from the pandemic context, social isolation measures, and maladaptive coping strategies.

## Data Availability

The datasets used and/or analysed during the current study are available from the corresponding author on reasonable request.

## Compliance with Ethical Standards

### Research involving Human Participants and/or Animals

All ethical guidelines concerning research involving human subjects were complied with and the Institutional Review Boards at the Universidade Nova de Lisboa and the University of São Paulo approved the research project before data collection began.

### Informed consent

All the participants signed free and informed consent forms online before proceeding with the questionnaire.

### Availability of data and materials

The datasets used and/or analyzed during this study are available from the corresponding author upon reasonable request.

